# Standard care vs. awake prone position in adult non-intubated patients with acute hypoxaemic respiratory failure secondary to COVID-19 infection – A multi-centre feasibility randomized controlled trial

**DOI:** 10.1101/2021.03.13.21253499

**Authors:** Devachandran Jayakumar, Pratheema Ramachandran, Ebenezer Rabindrarajan, Bharath Kumar Tirupakuzhi Vijayaraghavan, Nagarajan Ramakrishnan, Ramesh Venkataraman

## Abstract

**Background:** The primary manifestation of Corona Virus Disease −2019 (COVID-19) is acute hypoxic respiratory failure secondary to pneumonia and/or acute respiratory distress syndrome. Prone position has been shown to improve outcomes in ventilated patients with moderate to severe acute respiratory distress syndrome. The feasibility and safety of awake prone positioning and its impact on outcomes if any, in non-intubated patients with mild to moderate acute respiratory distress syndrome secondary to COVID-19 is unknown. Results of the observational studies published thus far in this pandemic have been conflicting. In this context, we conducted a multi-centre, parallel group, randomised controlled feasibility study on awake prone positioning in non-intubated patients with COVID-19 pneumonia requiring supplemental oxygen.

**Methods:** 60 patients diagnosed with acute hypoxic respiratory failure secondary to COVID −19 pneumonia requiring 4 or more litres of oxygen to maintain a saturation of ≥ 92% were recruited in this study. Thirty patients each were randomised to either standard care or awake prone group. Patients randomised to the standard care were allowed to change their position as per comfort and patients randomized to the prone group were encouraged to self-prone for at least 6 hours a day. The primary outcome was the proportion of patients adhering to the protocol in each group. Secondary outcomes include failure of therapy leading to escalation of respiratory support, number of hours prone, maximum hours of continuous prone positioning in a day, length of stay in ICU, ICU mortality, total number of patients needing intubation and adverse events.

**Results:** In the prone group, 43% (13 out of 30) of patients were able to self-prone for 6 or more hours a day. The median maximum prone duration per session was 2 hours. In the supine group, 47% (14 out of 30) were completely supine and 53% spent some hours in the prone position, but none exceeded 6 hours. There was no significant difference in any of the secondary outcomes between the two groups and there were no adverse events.

**Interpretation:** Awake proning in non-intubated patients with acute hypoxic respiratory failure is feasible and safe under clinical trial conditions. The results of our feasibility study will potentially help in the design of larger definitive trials to address this key knowledge gap.

## Background

Acute hypoxemic respiratory failure (AHRF) is one of the common causes of admission to intensive care units. Cardiogenic pulmonary oedema, pneumonia and acute respiratory distress syndrome (ARDS) account for the majority of cases of AHRF. The primary manifestation of Coronavirus Disease 2019 (COVID-19) caused by the Severe Acute Respiratory Syndrome Coronavirus-2 (SARS-CoV2) is AHRF secondary to pneumonia and/or ARDS.

ARDS is a rapidly progressive illness and is associated with substantial morbidity and mortality. Decades of research have not only improved our understanding of the pathophysiology of ARDS, but also helped identify treatments that improve survival. Two such interventions – lung protective ventilation and prone positioning have been identified in high-quality randomized trials to substantially reduce mortality^1,2^. Following on from these trials, prone ventilation has become an established therapy in ventilated patients with moderate to severe ARDS, although uptake and implementation of the intervention remains variable across ICUs^3^. Proning improves oxygenation through various mechanisms such as improved ventilation perfusion matching (V/Q), shape matching of the lungs within the chest wall and offloading the weight of the heart from the lungs^4,5^.

Although prone position has not been shown to be beneficial in ventilated patients with mild ARDS, case reports, case series and published cohort studies during this pandemic suggest that prone positioning even in non-ventilated patients with mild-moderate ARDS improves oxygenation and possibly prevents intubation ^6,7,8,9^. The mechanisms by which prone position improves oxygenation in non-ventilated patients are thought to be similar to those ventilated ^10,11^. However, all the published data thus far suffer from broadly the same limitations-absence of a control group, selection bias, residual and unmeasured confounding, and small sample sizes among others. Several randomized controlled trials evaluating this intervention have been registered, but none have been published so far^12^. While awake proning appears to be safe from available evidence, none of the studies were powered to identify potential adverse events. Additionally, prone positioning, by improving oxygenation, may provide a false sense of reassurance and potentially delay invasive ventilation and escalation of respiratory support^13^. Moreover, several fundamental questions such as, how long can a patient comfortably lie prone continuously, what prone duration is likely to offer clinical benefit, and whether the cumulative prone hours have an effect on outcome remain largely unanswered.

In this background, we conducted a multicentre feasibility randomized controlled trial of prone positioning in non-intubated patients with COVID-19 pneumonia, requiring supplemental oxygen.

## Methods

### Study Setting

This parallel group feasibility trial was conducted across 3 tertiary care hospitals in Chennai, India. At all of these sites, patients with COVID-19 pneumonia were managed in designated locations in the hospital and as per institutional protocols, patients needing more than 4l of oxygen were managed in areas capable of providing intensive care.

### Participants

Adults admitted to the intensive care unit with proven or suspected COVID-19 infection leading to hypoxic respiratory failure were screened for eligibility to participate in the trial.

#### Inclusion and exclusion criteria

Patients ≥18 years of age and requiring 4 or more litres per minute (LPM) of supplemental oxygen to maintain SpO2 ≥92% or if ABG was available, PaO2/FiO2 ratio between 100 and 300 mmHg (mild to moderate ARDS) with PaCO2 less than 45 mmHg were included. Patients with AHRF and haemodynamic shock requiring <0.1mcg/kg/min of nor-epinephrine were also considered for inclusion.

Patients below 18 years of age, pregnant women, patients with hemodynamic shock requiring norepinephrine ≥ 0.1 mcg/kg/min, any GCS <15, patients who needed immediate intubation in the opinion of the treating clinician and patients with absolute or relative contraindications to prone positioning (spinal instability secondary to severe rheumatoid arthritis, life threatening cardiac arrhythmias) were excluded.

### Intervention, Comparator and Trial procedures

Patients randomized to either group received oxygen via nasal prongs, face mask, non-rebreather mask, high flow nasal cannula (HFNC) or Non-invasive ventilation (NIV) as per treating clinician discretion.

In the intervention arm, patients were encouraged by bedside nurses to lie prone for a minimum of 6 hours in a day (cumulative). Additional pillows were provided for comfort or to facilitate prone position. Patients randomised to standard care were allowed to change their position as per their comfort (supine, semi sitting, sitting or lateral). If patients in the standard arm wished to lie prone for comfort, this was allowed. However, nurses and the treating team would not actively encourage prone positioning in this arm. Prone position sessions were considered significant and recorded, only if a session lasted more than 30 minutes in both arms.

Food and comfort breaks were planned while patients were supine. Oxygen flow and fraction of inspired oxygen (FiO2) was titrated to maintain a saturation of ≥92% in both arms at all times.

For face masks, up to 10 LPM oxygen flow rate was allowed. Since FiO2 varies with the patient’s inspiratory effort, approximate values were used to calculate FiO2 (5 LPM – 30%; 6 LPM 35%; 7 LPM 40%; 8 LPM 45%; 9 LPM 50% 10LPM 60%)^14^.

For patients on HFNC, flow was set at maximum permissible level on the device used or 60 liters per minute in case of blenders, and FiO2 adjusted to the goal (≥92%). When FiO2 was ≤ 40%, flow was weaned in quantums of 10LPM till total flow was 20LPM. When FiO2 was ≤ 30% with a flow of 20LPM, the patient was taken off HFNC and placed on an oxygen mask or nasal prongs.

If the patient was on NRBM, it was applied snugly over the face. With NRBM, FiO2 varies with the patient’s peak inspiratory flow and the leak around the mask. Since it is difficult to estimate the exact FiO2, approximate values were used based on oxygen flow in LPM (10, 12 and 15 LPM will approximately provide 60, 70 and 85% FiO2 respectively). Oxygen flow was adjusted to maintain a saturation of ≥92%. When the flow was ≤ 10LPM, the reservoir was folded and oxygen weaned off aiming for a saturation of ≥92%.

For patients on NIV, we recommended that sites use an oronasal interface and dual limb circuits to administer NIV using critical care ventilators. FiO2 and PEEP were titrated to maintain SpO2 ≥92%. Prone position while on NIV was achieved with multiple standard pillows and a C-pillow if required for the face and head to accommodate the mask.

The decisions to escalate respiratory support from the initial device to a device higher and the decision to intubate were left to the treating team.

#### Escalation of respiratory support

Patients on HFNC could be placed either on noninvasive ventilation (NIV) or intubated and mechanically ventilated; Patients on NRBM could be placed on HFNC, NIV or intubated depending on the clinical situation. Intermittent use of NIV along with Face mask, NRBM or HFNC was also considered as escalation of respiratory support.

#### Data collection

Data including patient demographics, APACHE II scores, height and weight as reported by patient or family, comorbidities (Diabetes, Hypertension, Coronary artery disease, Respiratory diseases like asthma, chronic obstructive pulmonary disease and interstitial lung disease, and Chronic kidney disease), initial oxygen delivery device, FiO2, PaO2/FiO2 ratios on admission, at 2 hours and twice daily (wherever available) were collected. Daily fluid balance was collected from the nursing chart. Total number of hours spent in prone position in a day (cumulative) and number of prone sessions and their duration were also recorded from the position chart. This protocol was followed for 7 days, or until escalation of respiratory support to the next level or patient improvement to discharge or death, whichever occurred first. Data on the administration of steroids, Remdesivir, Tocilizumab and Heparin/low molecular weight heparin were also collected. If a patient was unable to lie prone, reason for the inability to lie prone (neck pain, back ache, abdominal compression, breathlessness or claustrophobia) and adverse events like pressure ulcers, vomiting and nerve compression, if any, were recorded.

### Outcome measures

Since the trial was designed as a feasibility study, the primary outcome measure was the proportion of patients adhering to the protocol in each group. Secondary outcomes included the proportion of patients requiring escalation of respiratory support in either group, number of hours prone and maximum hours of continuous prone positioning in a day, length of stay in the ICU, ICU mortality, adverse events and reasons for not lying prone.

### Sample size

Since this was a feasibility study and there was limited data available on event rates at the time of designing the trial, we decided to enroll a total of 60 patients. Information from this trial will be helpful in determining sample sizes for a definitive trial.

## Design Details

### Randomization and allocation concealment

Patients were randomized in blocks of four using a computerized random number generator. Allocation was concealed using sealed opaque envelopes. Sites were not aware of block sizes. Because of the nature of the intervention, neither the participants nor the treating clinicians were blinded.

### Statistical analysis

Categorical variables are presented as proportions and analyzed using Chi Square Test; continuous variables are summarized as means and standard deviations or median and interquartile range based on distribution. Student t-test and paired t test was used to compare means as appropriate. All tests were two-tailed and p less than 0.05 was considered significant.

### Ethics and Informed consent

This study was approved by institutional ethics committee for biomedical research, Apollo Hospitals, Chennai (Approval No: AVH-C-S-005/05-20). Written informed consent was obtained from all the participants before enrollment.

## Results

68 patients were screened for eligibility over four months (screening data was available only for two centres) of which 60 patients consented to participate across three sites (Fig 1). 30 patients were randomised to each group. Baseline characteristics are presented in Table 1 and were comparable between the two groups. The primary outcome of adherence to protocol was 43% among the patients in the prone group (13 patients completed an average of at least 6 hours a day in prone position). In the supine group, 47% (14 out of 30) were completely supine and 53% spent some hours in the prone position, but none exceeded 6 hours (Fig 2). 70% of the patients were able to lie prone for 4 hours a day. The median maximum duration per session in the prone group was 2 hours. There was no significant difference in the cumulative fluid balance, length of stay, respiratory escalation, other medications use or mortality between the groups (Table 2). Four patients (13.3%) needed intubation in each group. Two patients (7.3%) in each group were discharged against medical advice to other hospitals (DAMA; Table 2). Two patients allocated to the prone group could not lie prone due to breathlessness. There were no adverse events from the positional therapy (Table 2).

**Table 1:**
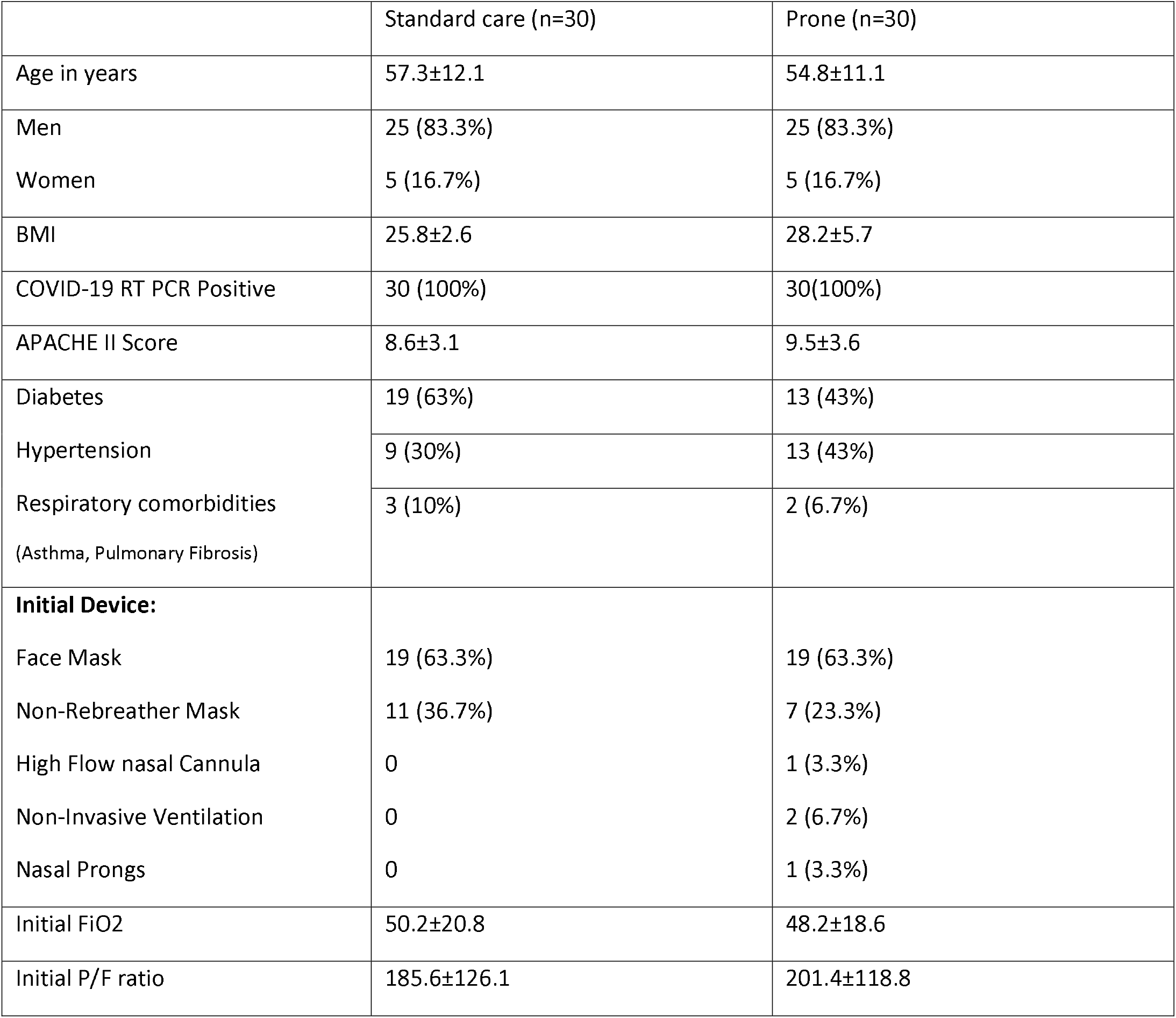
Baseline characteristics.

**Baseline characteristics: Table 1**
|  | Standard care (n=30) | Prone (n=30) |
| --- | --- | --- |
| Age in years | 57.3±12.1 | 54.8±11.1 |
| Men | 25 (83.3%) | 25 (83.3%) |
| Women | 5 (16.7%) | 5 (16.7%) |
| BMI | 25.8±2.6 | 28.2±5.7 |
| COVID-19 RT PCR Positive | 30 (100%) | 30(100%) |
| APACHE II Score | 8.6±3.1 | 9.5±3.6 |
| Diabetes | 19 (63%) | 13 (43%) |
| Hypertension | 9 (30%) | 13 (43%) |
| Respiratory comorbidities<br>(Asthma, Pulmonary Fibrosis) | 3 (10%) | 2 (6.7%) |
| **Initial Device:** |  |  |
| Face Mask | 19 (63.3%) | 19 (63.3%) |
| Non-Rebreather Mask | 11 (36.7%) | 7 (23.3%) |
| High Flow nasal Cannula | 0 | 1 (3.3%) |
| Non-Invasive Ventilation | 0 | 2 (6.7%) |
| Nasal Prongs | 0 | 1 (3.3%) |
| Initial FiO2 | 50.2±20.8 | 48.2±18.6 |
| Initial P/F ratio | 185.6±126.1 | 201.4±118.8 |

**Table 2:**
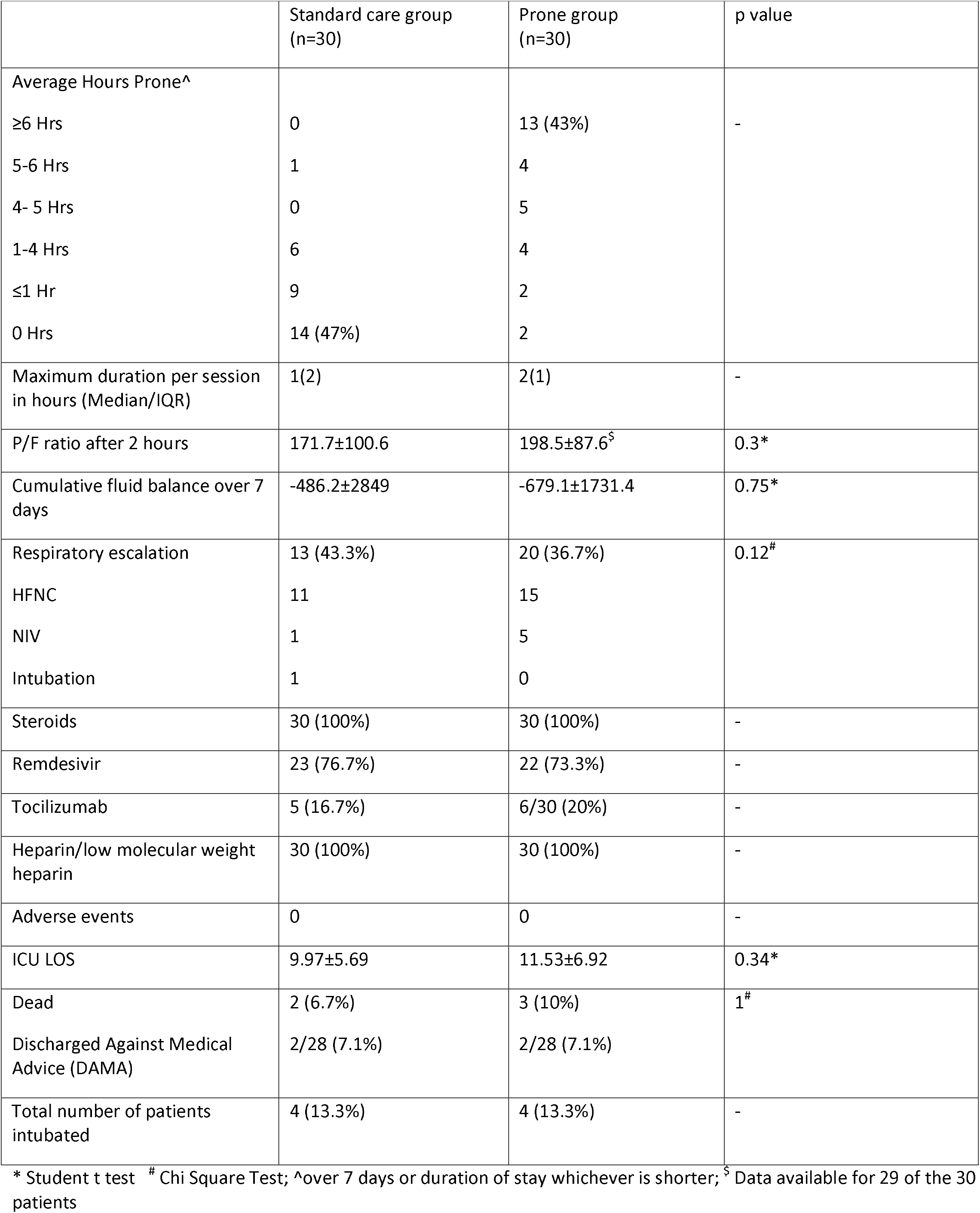
Primary and Secondary outcomes.

**Fig 1:**
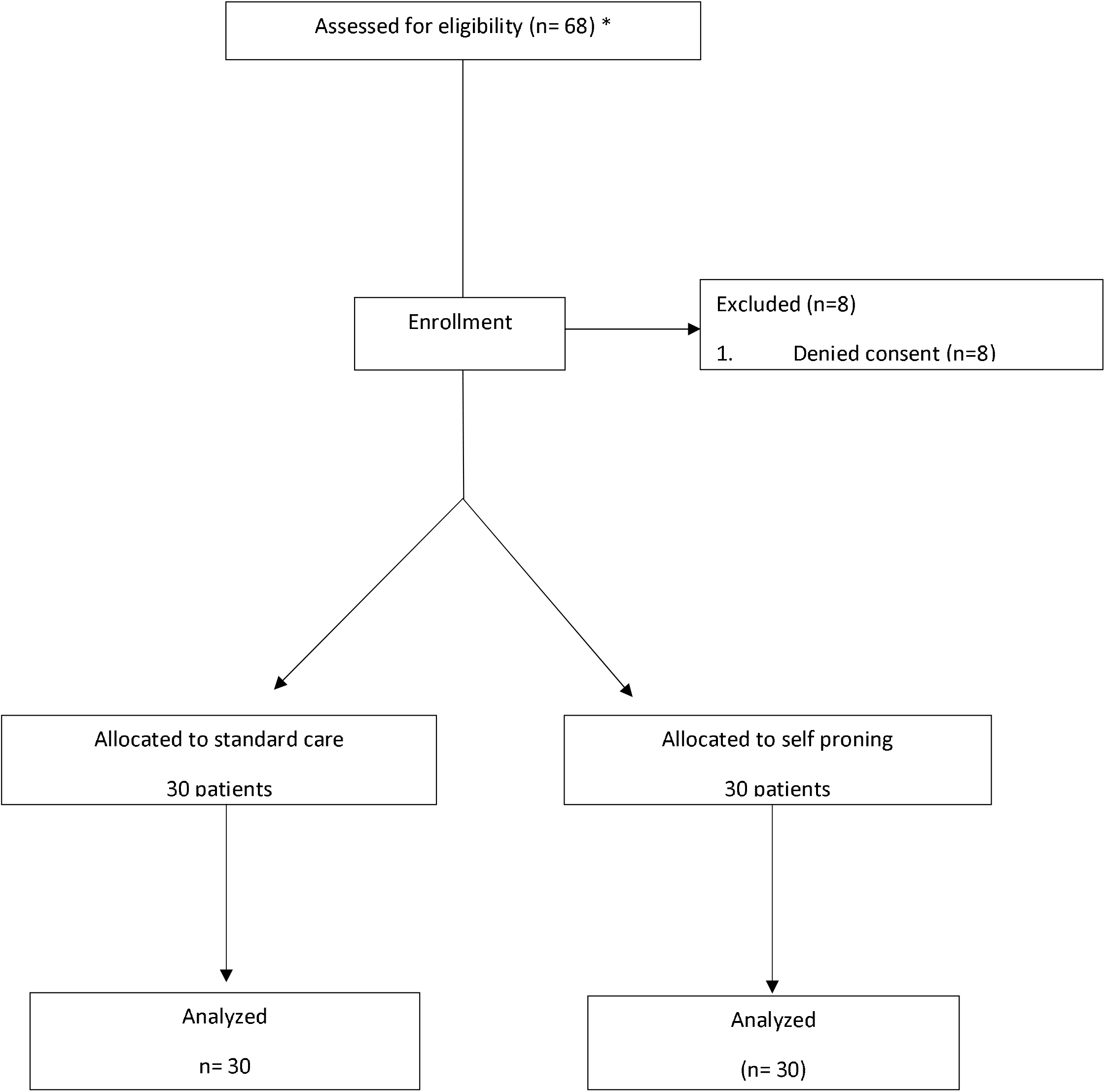
Consort Flow diagram. * Screening data available only for two centres

**Figure 2.**
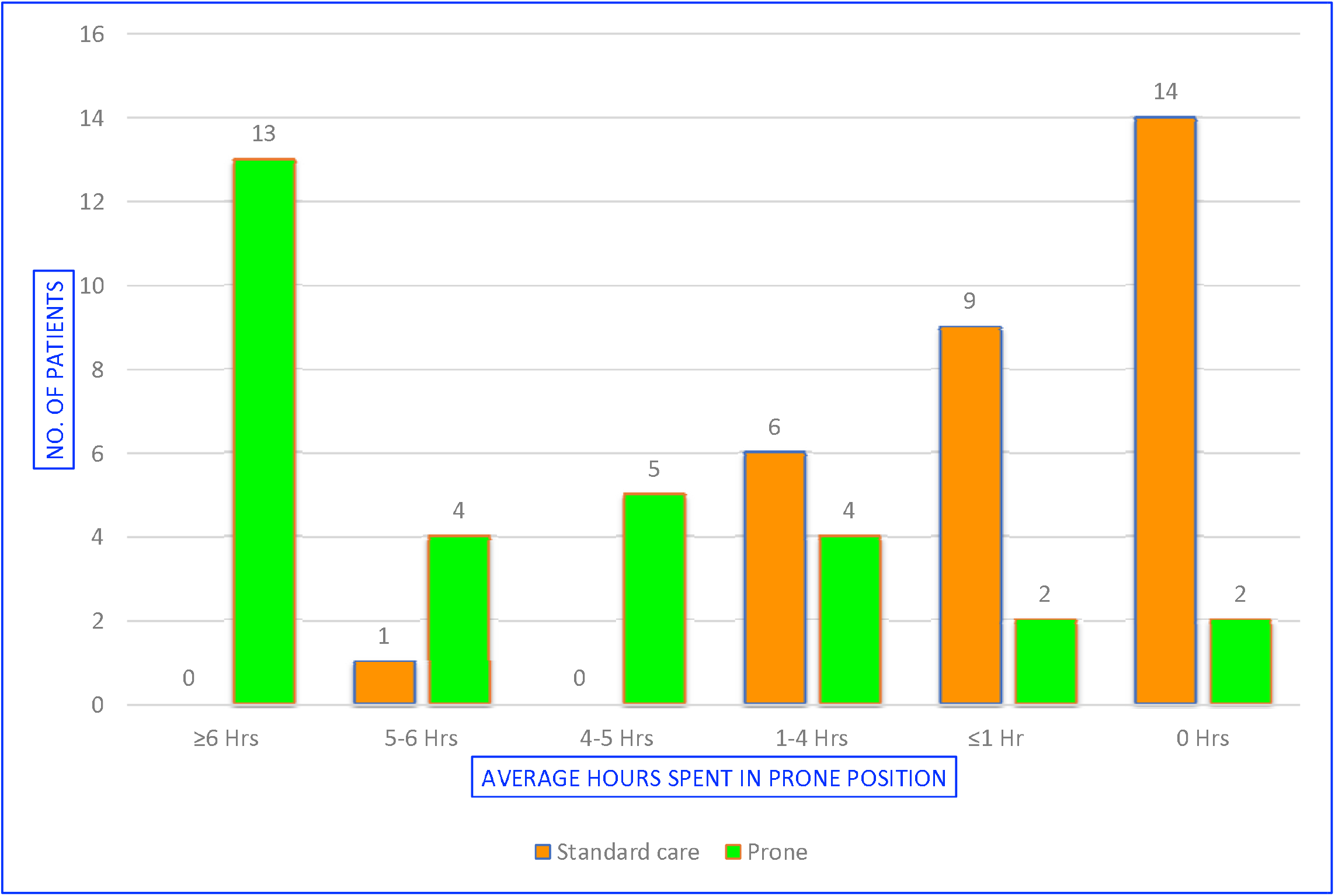

## Discussion

Our trial demonstrates the feasibility and safety of awake prone positioning for acute hypoxic respiratory failure secondary to COVID-19. Protocol compliance was 43% in the intervention arm. There was no difference in any of the secondary end points including the proportion of patients needing escalation of respiratory support, mortality or adverse events. Use of co-interventions was similar between the two groups.

While there are several clinical trials registered^12^, to our knowledge, this is the first randomized controlled trial evaluating awake proning for COVID-19 patients to be published. During the ongoing pandemic, there has been much interest in this intervention for a variety of reasons-the proven survival benefit of prone position in ventilated patients, the known favourable physiological benefits on oxygenation, the perceived ease of implementing the intervention, including outside an intensive care setting and the potential of the intervention in preventing escalation of respiratory support. These perceived advantages assume greater importance in healthcare systems and countries that are either by design, resource-constrained or in the face of the pandemic resource-overwhelmed.

In an observational single-centre study from France, Elharrar and colleagues included 24 patients with hypoxic respiratory failure from COVID-19 and evaluated the change in PaO2 (responders vs. non-responders) with prone position^7^. Responders were defined as those in whom there was an improvement of ≥20% in PaO2 during prone positioning. In their study, 63% of patients were able to lie prone for more than 3hours, 21% for 1-3hours and 4 patients who did not tolerate prone positioning for more than an hour. 6 patients (25% of the included cohort) were responders and among those that sustained prone postioning for 3hrs or more, the PaO2 improved from a mean of 73.6mm Hg (SD 15.9mmHg) to 94.9mmg Hg (SD 28.3mmHg).

In another single-centre cross-sectional study from Italy^8^, 15 patients receiving NIV in the prone position were included and changes in respiratory parameters were compared before, during and after prone positioning. Compared with the baseline, all included patients had a reduction in respiratory rate during and after prone positioning; all included patients also demonstrated an improvement in oxygen saturation and the PF ratio during prone positioning and 12 of the 15 patients had an improvement in the oxygen saturation and PF ratio after prone positioning. Similarly, Coppo and colleagues^10^ in a single-centre feasibility cohort study, demonstrated significant improvement in oxygenation from supine to prone position, but the effect did not sustain on upon resupination.

In contrast to the above studies, the PF ratio after 2hours was not different between the two arms in our trial. There are several possible reasons for this – lack of a true effect of the intervention, inadequate compliance (compliance with the intervention for 6hrs or longer was only 43%), improvements in oxygenation may be time dependent as well was related to the time since symptom onset (i.e., time from symptom onset to prone positioning) and differences in severity of illness.

While our trial and previous studies demonstrate safety, only a larger definitive trial can truly confirm this. As argued by Munshi and colleagues^15^, many factors influence the uptake of an intervention during a pandemic, including the perceptions of treatment risks and benefits, contextual factors such as ease of use and the characteristics of the physician (early or late adopters). While prone positioning appears to be a benign intervention, it is possible that the transient improvements in oxygenation may provide a false sense of assurance and in fact delay the escalation of respiratory support^13^. As such, the bar for accepting experimental interventions must not be lowered, even in the context of the desperation arising from the pandemic.

Our trial has several important strengths. We conducted a multi-centre feasibility evaluation of the intervention and employed appropriate strategies for randomization and allocation concealment. We collected data on key co-interventions and demonstrate no differences in the use of such treatments between the groups. Our trial now provides important information for the design of larger definitive trials in terms of feasibility, event rates and safety. The trial was conducted in centres at the peak of the pandemic when clinicians and hospital staff were coping with the enormous clinical burden and where clinical trial infrastructure was either nascent or absent. This is a key strength as it highlights the feasibility of such undertakings in countries and healthcare settings which are resource-constrained and typically excluded from such evaluations.

Our study also has important limitations. First, it was a feasibility study; therefore, the results are not powered to change practice. Second, it was not practically possible to collect the oxygenation data for every prone session; therefore, the protocol mandated a blood gas after two hours and twice daily blood gases thereafter to compute P/F ratios. This might not have accurately captured the improvement in oxygenation immediately after prone positioning. Third, only 43% could adhere to the protocol which required 6 hours of cumulative prone positioning in the prone group.

Several factors like change in nursing ratios, the overwhelming clinical burden, the need for isolation and cohorting which restricts access to trial personnel could have contributed to this low adherence, but it is important to note that 73% (22 out of 30) managed 4 or more hours of prone position a day. Whether the use of positional aids or mattresses will facilitate prolonged prone positioning is unknown and yet to be evaluated. Fourth, 53% (16 out of 30) in the supine group spent some hours in the prone position. Although this is a significant cross over, none of the patients exceeded 6 hours of prone positioning amounting to protocol violation. Fifth, the patients selected had mild to moderate illness severity and this explains the low mortality rate overall in both the groups. Sixth, onset of illness was not a criterion for inclusion. Some of these patients might have had illnesses for longer periods than others. This might have affected the overall efficacy of the intervention.

## Interpretation

Despite low adherence, our study confirms that awake prone positioning in non-intubated patients with AHRF is feasible and safe under clinical trial conditions and this data could potentially help construct protocols for future large randomised controlled trials. Future trials must include patients early, minimize cross over, improve prone positioning compliance to prolong the duration of sessions and could evaluate positional aids to prone.

## Registration

This feasibility trial is registered with Clinical Trials registry of India Registration number: CTRI/2020/12/029702.

## Data Availability

Deidentified data will be made available to authors upon request. Interested authors can contact the corresponding author by email for the data.

## Funding and approval

It’s an unfunded trial approved by the institutional ethics committee

## Authors’ Contributions

DJ – Conceptualisation, methodology, data curation, formal analysis, writing – original draft & editing.

PR – Project administration, supervision, and writing – review of the draft & editing.

ER – Project administration, resources, supervision and writing – review of the draft & editing.

BTV – Methodology, project administration, supervision, validation, writing – original draft, review & editing.

NR – Methodology, resources, supervision, validation and writing – review of the draft & editing.

RV – Methodology, project administration, supervision, validation, writing – original draft, writing – review & editing.

## Acknowledgements

We gratefully acknowledge the contributions of our colleagues Dr Babu K Abraham, Dr Senthil Kumar, Dr Ashwin K Mani, Dr Dedeepiya Devaprasad, Dr Vignesh Chandrasekaran, Dr Ashok Kumar, Dr Raymond Dominic Savio, Dr Ajay Padmanabhan, Dr Pavan Vecham, Dr Sristi Patodia, Dr Usha Rani and Dr Vanishree during the process of formulating the protocol and patient screening, and we thank our research team members Dr Manisha, Ms Evengeline Elvira, Ms Pavithra Sampathkumar and Mr Suresh Babu for their assistance in collecting and collating the data

